# Sex-specific neurodevelopmental outcomes in offspring of mothers with SARS-CoV-2 in pregnancy: an electronic health records cohort

**DOI:** 10.1101/2022.11.18.22282448

**Authors:** Andrea G. Edlow, Victor M. Castro, Lydia L. Shook, Sebastien Haneuse, Anjali J. Kaimal, Roy H. Perlis

## Abstract

**Importance:** Prior studies using large registries suggested a modest increase in risk for neurodevelopmental diagnoses among children of mothers with immune activation during pregnancy, and such risk may be sex-specific.

**Objective:** To determine whether in utero exposure to the novel coronavirus SARS-CoV-2 is associated with sex-specific risk for neurodevelopmental disorders up to 18 months after birth, compared to unexposed offspring born during or prior to the pandemic period.

**Design:** Retrospective cohort.

**Participants:** Live offspring of all mothers who delivered between March 2018 and May 2021 at any of eight hospitals across two health systems in Massachusetts.

**Exposure:** PCR evidence of maternal SARS-CoV-2 infection during pregnancy.

**Main Outcome and Measures:** Electronic health record documentation of ICD-10 diagnostic codes corresponding to neurodevelopmental disorders.

**Results:** The pandemic cohort included 18,323 live births, including 877 (4.8%) to individuals with SARS-CoV-2 positivity during pregnancy. The cohort included 1806 (9.9%) Asian individuals, 1634 (8.9%) Black individuals, 1711 (9.3%) individuals of another race, and 12,694 (69%) White individuals; 2614 (14%) were of Hispanic ethnicity. Mean maternal age was 33.0 years (IQR 30.0-36.0). In adjusted regression models accounting for race, ethnicity, insurance status, hospital type (academic center vs. community), maternal age, and preterm status, SARS-CoV-2 positivity was associated with statistically significant elevation in risk for neurodevelopmental diagnoses among male offspring (adjusted OR 1.99, 95% CI 1.19-3.34; p=0.009) but not female offspring (adjusted OR 0.90, 95% CI 0.43-1.88; p=0.8). Similar effects were identified using matched analyses in lieu of regression.

**Conclusion and Relevance:** SARS-CoV-2 exposure in utero was associated with greater magnitude of risk for neurodevelopmental diagnoses among male offspring in the 12 months following birth. As with prior studies of maternal infection, substantially larger cohorts and longer follow-up will be required to reliably estimate or refute risk.

**Trial Registration:** NA

**Key Points:** *Question:* Are rates of neurodevelopmental disorder diagnoses greater among male or female children with COVID-19 exposure in utero compared to those with no such exposure?

*Findings:* In a cohort of 18,323 infants delivered after February 2020, males but not females born to mothers with a positive SARS-CoV-2 PCR test during pregnancy were more likely to receive a neurodevelopmental diagnosis in the first 12 months after delivery, even after accounting for preterm delivery.

*Meaning:* These findings suggest that male offspring exposed to COVID-19 in utero may be at increased risk for neurodevelopmental disorders.

## Introduction

Multiple epidemiologic studies using large-scale administrative or health registry data sets suggest that maternal infection or inflammation during pregnancy is associated with risk for a range of neurodevelopmental disorders among offspring. Two early studies – one using data from Helsinki, Sweden^1^, another using national data from Denmark^2^ – both identified association between pregnancy during periods of influenza exposure and schizophrenia hospitalization. A study of similar design using UK data from the 1957 influenza pandemic likewise identified elevated rates of schizophrenia^3^. However, all three studies relied on birth dates relative to pandemic activity, rather than examining documented maternal infection, raising the possibility that some other aspect of pregnancy during the pandemic period could confer risk. For example, such pandemics might be associated with dietary changes or greater overall levels of maternal stress exposure.

More recent investigations have directly examined the association with infection outside of pandemic periods. A Swedish study of 2.3 million births found a 30% increase in risk for autism among offspring of women with an inpatient infection diagnosis during pregnancy^4^. A subsequent analysis of 1.8 million births in Sweden identified a 79% increase in risk for autism diagnosis in offspring, and a 24% increase in risk of major depression, following any maternal infection, without evidence of an effect of infection severity^5^. Animal model studies have demonstrated that viral and bacterial infection in pregnancy, as well as non-infectious maternal immune activation, are associated with a range of neurodevelopmental morbidities in offspring (reviewed in Shook^6^, Careaga^7^, and Meyer^8^).

In light of the magnitude of the COVID-19 pandemic, there is a critical public health need to understand the extent to which maternal exposure may have similar effects on offspring to those observed in these prior studies of infection in pregnancy. Findings of neuropsychiatric sequelae of SARS-CoV-2 infection among both adults and children raise concern that, even absent direct infection of the central nervous system, SARS-CoV-2 may exert persistent effects on the brain^9,10,11,12,13,14^.

Studies of maternal and placental immune response also suggest sex-specific responses to SARS-CoV-2 infection, with upregulation of placental interferon signaling and reduced maternal anti-SARS-CoV-2 antibodies among male compared to female pregnancies^15^. Sex-specific responses to maternal exposures have been posited to underlie well-established sex differences in the prevalence of neurodevelopmental and neuropsychiatric disorders like autism, attention deficit hyperactivity disorder, anxiety and major depression.^16,17,18^

Detecting neurodevelopmental effects of similar magnitude to those reported by prior studies will require large cohorts with long-term follow-up. Notably, small sample sizes and short-term follow-up surveys would not have identified the now-accepted neurodevelopmental risks associated with influenza or other maternal infections, at the magnitude commonly reported (e.g., hazards or odds ratios of 1.3-1.6 for autism after maternal infection in pregnancy). Electronic health record (EHR)-based studies are one way to rapidly assemble larger cohorts to address the urgent need for information about potential neurodevelopmental consequences of maternal SARS-CoV-2 infection. We previously reported preliminary evidence of an association between in utero exposure to SARS-CoV-2 and a delay in motor and speech milestones in children at one year of age^19^ based on electronic health record data. Here, we sought to examine this risk in a cohort four times larger, in order to address two questions that could not be explored in the prior study. First, are there sex-specific differences in risk, as might be predicted based on evidence of sexual dimorphism in COVID response and greater male vulnerability to neurodevelopmental insults? And second, does pregnancy during the pandemic itself independent of COVID-19 exposure, compared to pregnancy prior to the pandemic, confer neurodevelopmental risk?

## Methods

### Study design and data set generation

We applied the same cohort definitions as those previously reported^19^. Specifically, we queried electronic health records as represented in the electronic data warehouse spanning a total of 8 hospitals in Eastern Massachusetts, including 2 tertiary care medical centers and 6 community hospitals, as well as all affiliated outpatient networks. Data were extracted for Massachusetts General Hospital (MGH), Brigham and Women’s Hospital (BWH), Newton-Wellesley Hospital (NWH), North Shore Medical Center (NSMC), Martha’s Vineyard Hospital (MVH), Nantucket Cottage Hospital (NCH), Cooley Dickinson Hospital (CDH), and Wentworth Douglass Hospital (WDH), which share a common electronic data warehouse and governance.

We began by identifying every live birth in one of these hospitals beginning March 1, 2020, and ending May 31, 2021, representing the pandemic cohort. We generated a new comparison cohort not incorporated in our prior investigation, including all live births between January 1 and December 31, 2018, representing a cohort of children born and followed up prior to the pandemic. For further sensitivity analysis, we also examined a third cohort born prior to the pandemic, but followed during the pandemic at least in part, spanning all live births between January 1 and December 31, 2019, to evaluate for ascertainment bias that might make physicians more likely to diagnose neurodevelopmental disorders in children during the pandemic. For all cohorts, we linked offspring to pregnant parent based upon medical record number, date and time of birth, and offspring sex. We characterized medical history for the birthing person via ICD-10 billing codes, problem lists, medications, and laboratory studies occurring between the date of estimated last menstrual period and the discharge date for the delivery admission. COVID-19 vaccination status was drawn from documentation in the electronic data warehouse, which integrates vaccination orders, reports of vaccine status, and regional public health records. Electronic health records were also used to determine sociodemographic features (maternal age, self-reported gender, insurance type, and self-reported race and ethnicity based on US Census categories), in order to facilitate control for potential confounding variables. The Massachusetts General Brigham Institutional Review Board approved all aspects of this study, with a waiver of informed consent as no patient contact was required, the study was considered to be minimal risk, and consent could not feasibly be obtained. We followed the STROBE reporting guideline for cohort studies.

### Outcome definition

For all offspring, we extracted ICD-10 billing codes and problem lists. The primary outcome was defined as any diagnosis of a neurodevelopmental disorder, based on presence of at least one ICD10 code included in the Healthcare Cost and Utilization Project (HCUP) level 2 ‘developmental’ category (code 654). These codes included F8x (pervasive and specific developmental disorders: developmental disorders of speech and language [F80]; specific developmental disorders of scholastic skills [F81]; specific developmental disorder of motor function [F82]; pervasive developmental disorders [F84]; other/unspecific disorder of psychological development [F88/89]) and F7x (intellectual disabilities). As in our prior work, we allowed diagnoses that are not typically assigned in the first 18 months of life, to allow for a consistent definition that could be preserved across age groups in future investigations.

### Exposure definition

The electronic data warehouse integrates laboratory SARS-CoV-2 PCR results from hospital network laboratories as well as laboratories outside the network. Every birth was linked to the pregnant individual’s test results at any point during pregnancy. Universal screening for SARS-CoV-2 by PCR on admission to Labor and Delivery was implemented across the Massachusetts General – Brigham (MGB) health systems in April 2020 and continues to the present. Thus, at minimum, asymptomatic pregnant patients were tested at the time of hospitalization. During the study period, patients with positive home rapid antigen testing were advised to confirm/document a positive home test with hospital-accessible PCR, per hospital infection control policy. Those individuals with only negative PCR results documented were considered to be negative.

### Analysis

We fit logistic regression models among males and females examining the association between maternal SARS-CoV-2 status and presence or absence of neurodevelopmental outcome at any point within the first 12 months following birth, adjusting for maternal age in years, race and ethnicity, insurance type (public versus private), and hospital type, to yield adjusted estimates of effect and 95% confidence intervals. Survival analysis was not applied as 12 months of follow-up was available for all offspring. Primary models also included preterm delivery status, recognized as a risk factor for neurodevelopmental outcomes that has been associated repeatedly with SARS-CoV-2 positivity during pregnancy. Thus, primary models sought to examine risk associated with SARS-CoV-2 positivity, after accounting for preterm delivery as well as potentially confounding sociodemographic features. In a pooled analysis, we also included an indicator variable for offspring sex. Non-independence of multiple births was addressed by considering observations to be clustered within deliveries; the glm.cluster command in the R miceadds package (v3.11-6) to generate robust standard errors. As 18-month outcomes were available for a smaller subset, we secondarily explored outcomes among this group.

To examine the robustness of multiple regression results by applying an alternative approach to potential confounding variables, we also applied exact matching to match each COVID-exposed offspring with at least one COVID-unexposed offspring on the same features used in regression models (i.e., race, ethnicity, maternal age, insurance type, hospital type, and preterm delivery). Where regression models estimate an average treatment effect, the matched cohort provides an approximation of the average treatment (i.e., exposure) effect among exposed individuals. We used the R matchit (v4.4.0) package for these analyses, allowing as many exact matches among controls as available – i.e., 1:n matching. Repeating matched analyses using coarsened exact matching yielded essentially the same results.

Finally, to test the hypothesis that there might be greater rates of neurodevelopmental diagnoses among offspring of pandemic-era pregnancies, we compared this pandemic-born birth cohort (regardless of SARS-CoV-2 exposure status) to two different groups. The first included all live births in 2018, allowing for a full 12 months of follow-up prior to the pandemic. The second included all live births in 2019, recognizing that some follow-up would occur during the pandemic which might itself impact detection of neurodevelopmental outcomes. As in primary analyses, we stratified by sex, then pooled males and females. All analyses used R 4.0.3 (The R Foundation for Statistical Computing, Vienna, Austria), with statistical significance defined as uncorrected two-tailed p<0.05.

## Results

The pandemic cohort included 18,323 live births, including 877 (4.8%) to mothers with SARS-CoV-2 positivity during pregnancy; the latter group was more likely to be Hispanic, of non-White race, and to have public versus private insurance (Table 1). The full cohort of mothers included 1806 (9.9%) Asian individuals, 1634 (8.9%) Black individuals, 1711 (9.3%) individuals of another race, and 12,694 (69%) White individuals; 2614 (14%) were of Hispanic ethnicity. Mean maternal age was 33.0 years (IQR 30.0-36.0). Of the 877 COVID-exposed offspring, 26 (3.0%) received a neurodevelopmental diagnosis during the first 12 months of life (this included 0 of 13 whose mothers were partially or fully vaccinated at time of infection), as compared to 316 (1.8%) among the COVID-unexposed offspring; specific neurodevelopmental diagnostic codes are listed in Supplemental Table 1. Three SARS-CoV-2 unexposed children, and none of the exposed children, died prior to 12 months of follow-up.

**Table 1.** Socioemographic and baseline clinical characteristics of live births to mothers who were SARS-CoV-2 positive or negative (March 2020 - May 2021).

| Characteristic | Pregnancy<br>SARS-COV-2 Negative,<br>N = 17,446 | Pregnancy<br>SARS-COV-2<br>Positive, N = 877 | p-value<sup>2</sup> |
| --- | --- | --- | --- |
| **Maternal age, Mean (SD)** | 33.1 (4.8) | 31.3 (5.6) | <0.001 |
| **Maternal race, n (%)** |  |  | <0.001 |
| Asian | 1,760 (10) | 46 (5.2) |  |
| Black or African American | 1,495 (8.6) | 139 (16) |  |
| Other | 1,475 (8.5) | 236 (27) |  |
| Unknown | 428 (2.5) | 50 (5.7) |  |
| White | 12,288 (70) | 406 (46) |  |
| **Maternal ethnicity, n (%)** |  |  | <0.001 |
| Hispanic | 2,279 (13) | 335 (38) |  |
| Not Hispanic | 14,677 (84) | 508 (58) |  |
| Unavailable | 490 (2.8) | 34 (3.9) |  |
| **Maternal public insurance, n (%)** | 3,162 (18) | 427 (49) | <0.001 |
| **Trimester of maternal SARS-COV-2 infection, n (%)<sup>\*</sup>** |  |  | >0.99 |
| 1st | 0 (NA) | 62 (7.4) |  |
| 2nd | 0 (NA) | 233 (28) |  |
| 3rd | 0 (NA) | 546 (65) |  |
| **Delivery hospital type, n (%)** |  |  | <0.001 |
| Academic medical center | 10,167 (58) | 622 (71) |  |
| Community hospital | 7,279 (42) | 255 (29) |  |
| **Delivery method, n (%)** |  |  | 0.46 |
| C-Section | 5,719 (33) | 298 (34) |  |
| Vaginal | 11,727 (67) | 579 (66) |  |
| **Multiple births, n (%)** | 1,094 (6.3) | 71 (8.1) | 0.031 |
| **Delivery admission length of stay (days), Median (IQR)** | 3.00 (2.00 – 4.00) | 3.00 (2.00 – 4.00) | 0.62 |
| **Pre-term birth, n (%)** |  |  | 0.001 |
| Preterm | 1,736 (10.0) | 117 (13) |  |
| Term | 15,710 (90) | 760 (87) |  |
| **Offspring gender, n (%)** |  |  | 0.59 |
| Female | 8,518 (49) | 420 (48) |  |
| Male | 8,928 (51) | 457 (52) |  |

<sup>1</sup>Wilcoxon rank sum test; Pearson's Chi-squared test; Fisher's exact test. Other race includes American Indian or Alaska Native, Native Hawaiian or Other Pacific Islander
\* Trimester unknown for n=36

In regression models adjusted for race, ethnicity, insurance status, hospital type, maternal age, and preterm status, statistically significant elevation in risk associated with maternal SARS-CoV-2 positivity was observed among male offspring (adjusted OR 1.99, 95% CI 1.19-3.34; p=0.009; Figure 1A) but not female offspring (adjusted OR 0.90, 95% CI 0.43-1.88; p=0.8; Figure 1B). In pooled analysis, maternal SARS-CoV-2 positivity was associated with numerically but not statistically significantly greater rates of neurodevelopmental diagnoses (adjusted OR 1.45, 95% CI 0.95-2.21; p=0.09; Figure 1).

**Figure 1A.**
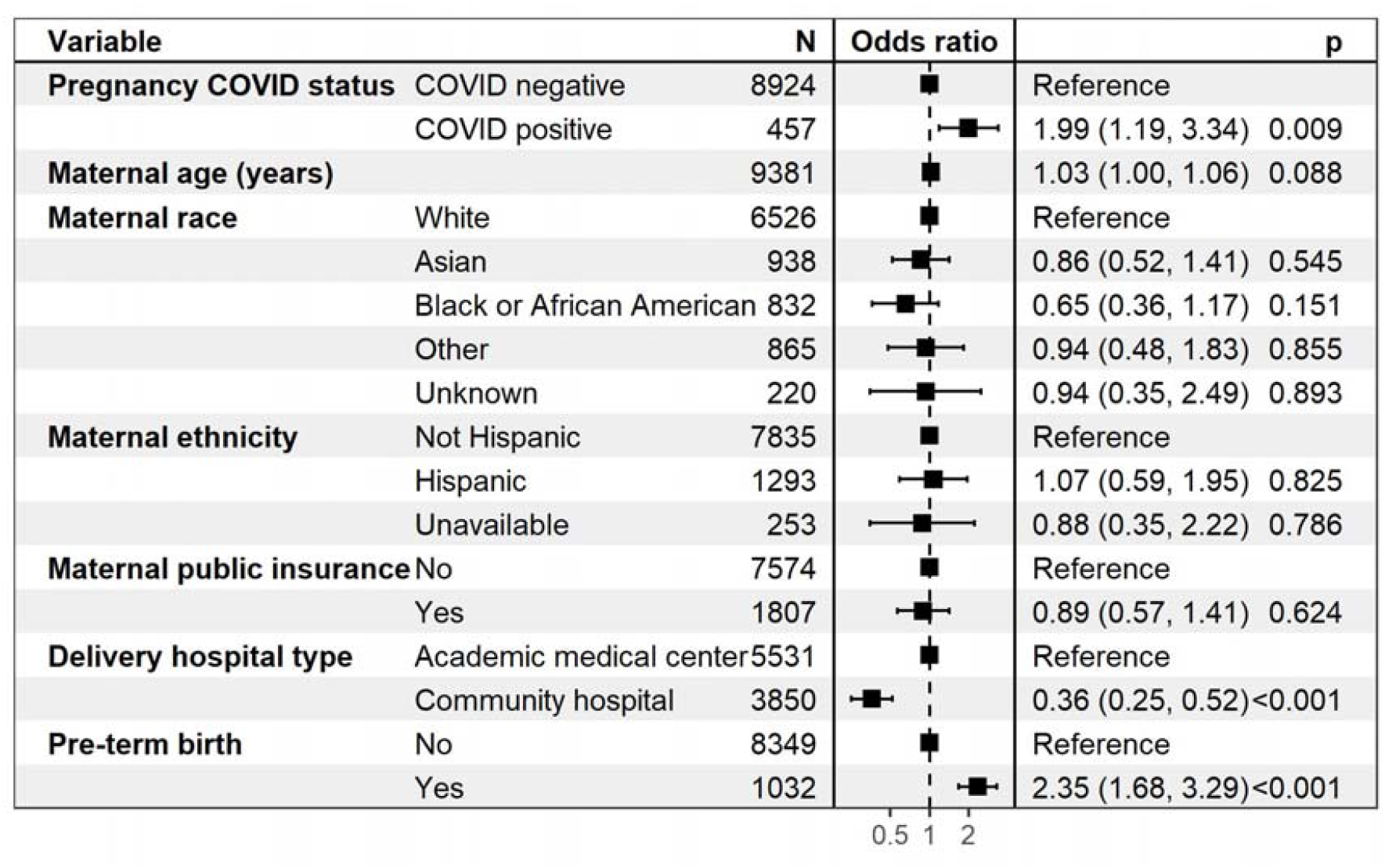
Logistic regression forest plot of neurodevelopmental disorder outcome up to 12 months in male offspring stratified by maternal COVID-19 status, maternal age, maternal race and ethnicity, maternal insurance type, delivery hospital type, and pre-term birth.

**Figure 1B.**
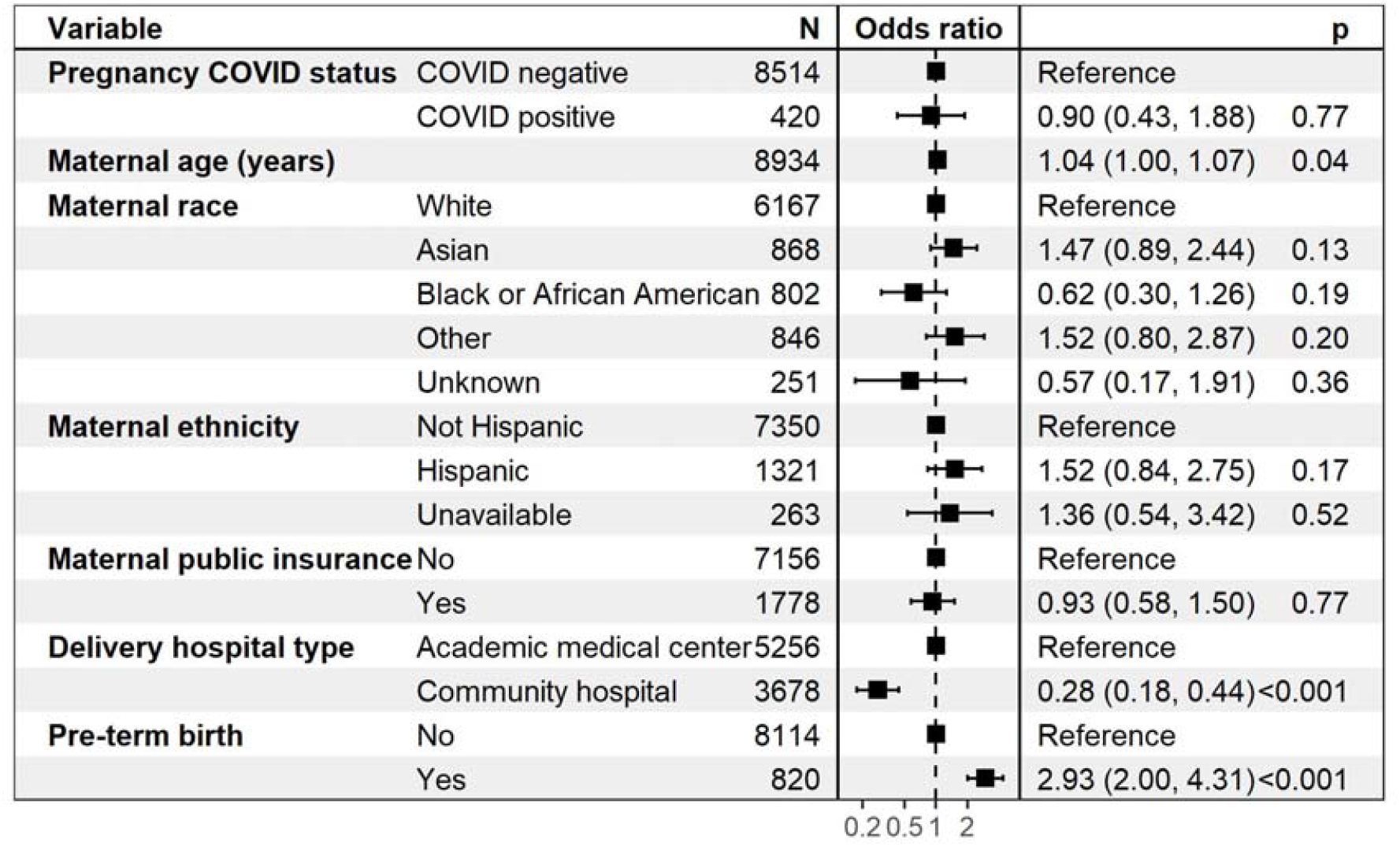
Logistic regression forest plot of neurodevelopmental disorder outcome up to 12 months in female offspring stratified by maternal COVID-19 status, maternal age, maternal race and ethnicity, maternal insurance type, delivery hospital type, and pre-term birth.

**Figure 1C.**
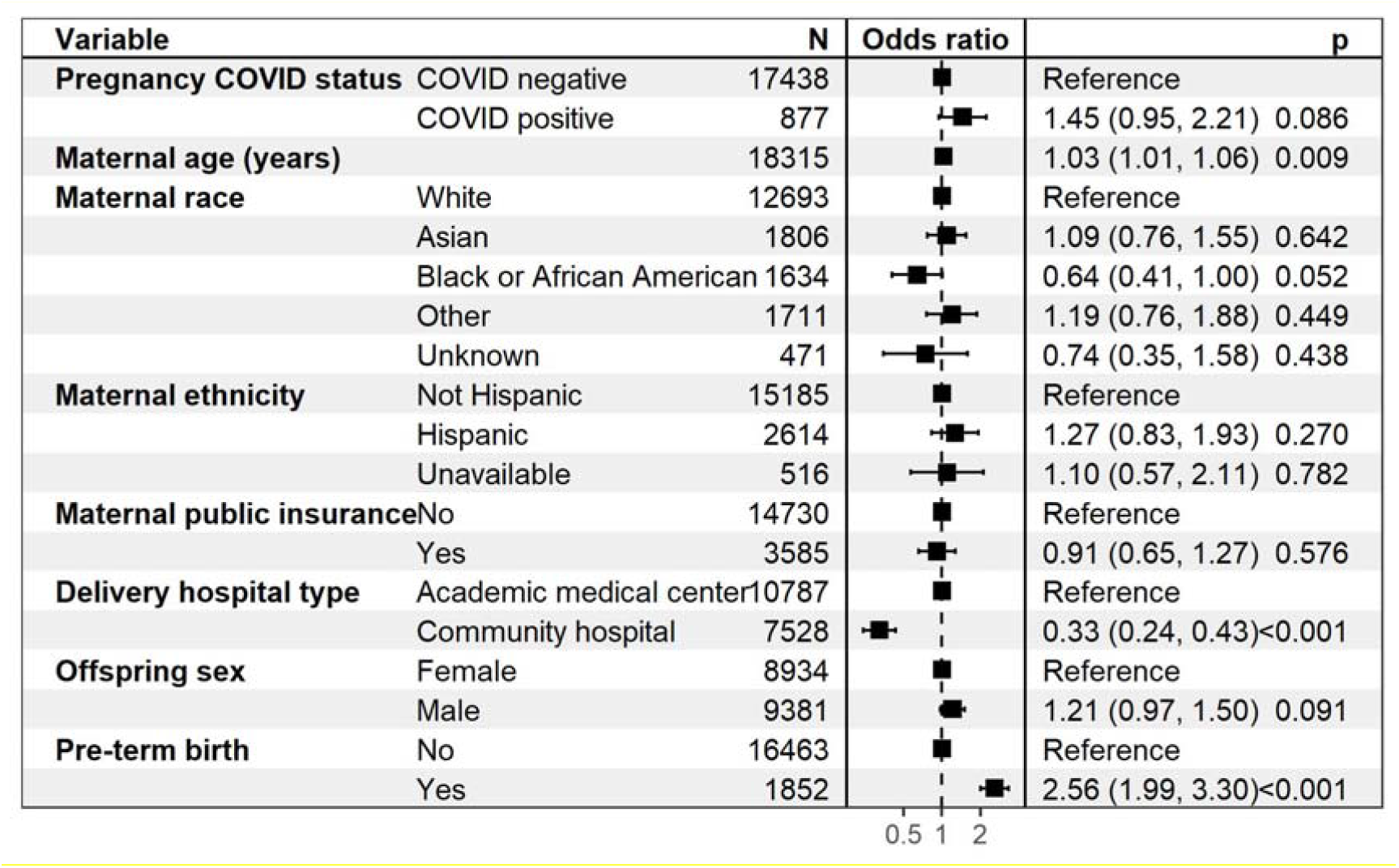
Logistic regression forest plot of neurodevelopmental disorder outcome up to 12 months in all offspring stratified by maternal COVID-19 status, maternal age, maternal race and ethnicity, maternal insurance type, delivery hospital type, and pre-term birth.

As an examination of the robustness of these effects, we repeated these analyses using exact matching on the same features as regression – maternal age, race, ethnicity, insurance status; hospital type; and preterm status. Among males, matching 394 COVID-exposed offspring with 6014 COVID-unexposed offspring, OR was 1.88 (95% CI 1.05-3.37; p=.03; Supplemental Figure 1A), and for female offspring matching 379 COVID-exposed to 5792 COVID-unexposed, OR was 1.22 (0.56-2.65, p=.6; Supplemental Figure 1B). For the cohort as a whole, matching 773 COVID-exposed offspring with 11806 COVID-unexposed offspring, OR was 1.58 (95% CI 0.99-2.52; p=.05; Supplemental Figure 1C). These sensitivity analyses demonstrated consistent direction of effect across both matched analyses and logistic regression models. Similarly, repeating regression analyses *without* adjustment for pre-term birth (Supplemental Figures 3A, B, and C) yielded similar results: adjusted OR 2.05 (95% CI 1.22-3.43, p=0.006), 1.04 (95% CI 0.45-1.99, p=.89), and 1.55 (95% CI 0.99-2.30, p=.06) for male, female, and pooled offspring, respectively.

Outcomes at 18 months were available for 13435 COVID-unexposed and 551 COVID-exposed offspring (Supplemental Table 2). As with 12-month outcomes, greater numeric magnitude of risk was observed among male offspring (adjusted OR 1.44, 95% CI 0.96-2.18, p=.08) but not female offspring (adjusted OR 0.99, 95% CI 0.55-1.78, p=.96). In pooled analysis, COVID exposure was associated with a numeric but not statistically significant increase in neurodevelopmental outcomes (OR 1.27, 95% CI 0.91-1.77; p=.165). Omitting preterm delivery as a covariate again yielded similar results (Supplemental Figures 4A, B, and C.)

Secondarily, we also tested for secular trends in diagnosis of neurodevelopmental disorders by comparing 12-month outcomes among children born during the pandemic to two pre-pandemic cohorts. Compared to 2018 (Supplemental Table 3), a modest but not statistically significant increase in rates of neurodevelopmental diagnoses (Supplemental Table 5) was identified among males (adjusted OR 1.10, 95% CI 0.88-1.37), p=.41) as well as females (adjusted OR 1.25, 95% CI 0.96-1.62), p=.09); pooling both sexes, adjusted OR=1.16, 95% CI 0.98-1.37, p=.08)(Supplemental Figures 5A, B, and C). Compared to 2019 (Supplemental Table 4), adjusted OR for male offspring was 1.24, 95% CI 0.99-1.55, p=.06; for females, 1.22, 95% CI 0.95-1.57, p=.12; and pooled, 1.23, 95% CI 1.04-1.46, p=.015; Supplemental Figures 6A, B, and C).

## Discussion

Among a cohort of 18,323 deliveries during the pandemic, we identified a statistically significant elevation in risk solely among male but not female offspring, detectable using two complementary approaches to address potential confounding variables. These effects were not attributable to preterm delivery, which was significantly more common among COVID-exposed offspring. As all deliveries occurred during the pandemic period, these effects also cannot be attributed to nonspecific effects of pandemic-era stress. We also detected an increase in risk for neurodevelopmental diagnoses, more modest in magnitude and similar among male and female offspring, associated with pandemic-era deliveries compared to 2018 and 2019; these latter effects were statistically significant only in a pooled analysis of both sexes for 2019.

In general, our results are consistent with abundant evidence that exposure to infection during pregnancy – including viral infections such as influenza – is associated with an increased risk for neurodevelopmental morbidity in offspring^5,20–24^. Such risk was initially detected as an increase in schizophrenia and autism diagnoses following influenza and rubella pandemics^123^, was recapitulated in animal models^25268^, and was more recently directly tested in large registry studies^4,5^. Risk of neurodevelopmental disorders in offspring after maternal prenatal exposures also has been reported to be greater in male offspring^27,28^. Because the neurodevelopmental risk in offspring is thought to be mediated in large part through maternal and placental immune activation (reviewed in Shook^6^, Careaga^7^, and Meyer^8^), and has been observed in other viral infections that, similar to SARS-CoV-2, are not thought to directly infect fetal brain tissue^6^, it is biologically plausible that SARS-CoV-2 infection in pregnancy could impact offspring risk for neurodevelopmental disorders. Indeed, several studies demonstrated early signals (e.g. at 3-6 months, 12, and 18 months) of neurodevelopmental risk in offspring exposed to SARS-CoV-2 in utero^29,30(p19),31^, although those studies were limited by the absence of a contemporaneous non-infected control group. A preliminary report in a subset of the present cohort, including a non-infected contemporary control group, suggested that SARS-CoV-2 was associated with increased risk for neurodevelopmental diagnosis at 12 months of age,^19^ but did not yet have sufficient neurodevelopmental diagnoses to examine the impact of fetal sex, as we have now been able to examine in this substantially larger cohort.

This observation may still represent type 1 error, and further study will be required to more precisely understand the magnitude of risk that exists, if any. It is, however, consistent with abundant evidence that the developing male brain is more vulnerable to in utero environmental effects^32,33^, including immune activation^34^. In the case of SARS-CoV-2, prior work identified robust immune activation at the maternal-fetal interface^35,36^, and sexually dimorphic effects of maternal SARS-CoV-2 on placental immune response, with significant upregulation of Type I, II and III interferon signaling pathways in the placentas of pregnancies with male fetuses and downregulation in pregnancies with female fetuses.

While our cohort is as yet too small to evaluate whether there are protective effects of maternal vaccination on offspring neurodevelopmental risk if SARS-CoV-2 infection does occur in pregnancy, we do note that none of the neurodevelopmental outcomes occurred among COVID-exposed offspring of vaccinated mothers. There are abundant benefits associated with vaccination for pregnant women^37(p19)^. If confirmed in larger samples, our results may suggest additional benefit, namely mitigating some of the potential risk associated with SARS-CoV-2 infection during pregnancy. As with our other analyses, additional investigation will be required to exclude the potential for confounding in this context as well.

We detected a more modest elevation in risk associated with birth during the pandemic period itself. We emphasize that such an effect cannot explain the COVID associations we observe, as primary analyses are limited to the pandemic period. There are instead a wealth of potential explanations for such a secular trend, many unrelated to biology. While greater maternal stress could represent one such explanation, as posited by a recent small study with similar findings^38^, absent direct measures of stress, this conclusion would be purely speculative. Other potential explanations for this finding that are unrelated to maternal biology could include ascertainment bias (e.g., providers being more likely to diagnose neurodevelopmental disorders in children born during the pandemic), changes in social environment for offspring during the pandemic, or even changes in billing as health systems adapt to remote assessment. These alternate explanations merit investigation in future studies.

## Limitations

While the present study expands the size of our cohort 4-fold, we emphasize the importance of larger scale investigation with longer-term follow-up to understand the potential neurodevelopmental risks, and to investigate key questions related to trimester of exposure and vaccination status. It is not clear that the changes we can detect at 12 and 18 months will be indicative of persistent risks for disorders such as autism, intellectual disability, or schizophrenia. For this reason, systematic prospective neuropsychiatric phenotyping in children with in utero exposure will be particularly important to complement and extend our results.

Some of the potential biases due to missing data could tend to increase the likelihood of a null finding. For example, some mothers with no documented SARS-CoV-2 infection could actually have had asymptomatic infection, or an inability to access testing during illness. While universal screening and the standardized infection control practices referenced should ensure accuracy of the record for SARS-CoV-2 infection in pregnancy, it is certainly possible that some patients with only a negative PCR result available at delivery were positive at an earlier point in pregnancy that was not captured with a PCR test; such undetected positivity among our “control” population would be more likely to bias our results toward the null.

Children who were evaluated outside of the outpatient pediatric networks of these 8 hospitals could have received neurodevelopmental diagnoses not documented in follow-up pediatric notes. In the case of neurodevelopmental outcomes, results could be biased if mothers with COVID-19 during pregnancy receive closer follow-up, increasing the likelihood of detection of any diagnosis in this group. In our prior work, we have shown that other diagnoses are not inflated, as would be expected if children of previously-ill mothers were more likely to be evaluated or receive more frequent pediatric assessment^19^. For further discussion of collider bias as it relates to COVID-19, see Griffith^39^. Nonetheless, these limitations underscore the importance of multiple, complementary follow-up study designs, rather than reliance on any single investigation or methodology.

## Conclusion

Despite these limitations, this cohort study supports the hypothesis that male children exposed to maternal SARS-CoV-2 infection in utero may be at modestly greater risk for adverse neurodevelopmental outcomes. This study further suggests a small secular trend of increased neurodevelopmental diagnoses in children born intrapandemic compared to pre-pandemic births, independent of infection status. This too, could reflect a type of ascertainment bias with providers more likely to be attuned to neurodevelopmental diagnoses in children born during the pandemic. In aggregate, our results underscore the need for larger-scale studies applying a range of follow-up strategies, to more precisely estimate and characterize risk.

## Data Availability

Data are not available because of restrictions on use of electronic health records.

## Acknowledgements

This study was supported by the National Institute of Mental Health (RF1MH132336, Dr. Edlow and Dr. Perlis; R01 MH116270, Dr. Perlis; U54 MH118919, Dr. Edlow), the National Institute of Child Health and Human Development (R01 HD100022-02S2, Dr. Edlow), and Simons Foundation SFARI grant 870754 (Dr. Edlow). The sponsors did not contribute to any aspect of the design and conduct of the study; collection, management, analysis, and interpretation of the data; preparation, review, or approval of the manuscript; and decision to submit the manuscript for publication. The authors had the final responsibility for the decision to submit for publication. Dr. Perlis and Edlow had full access to all the data in the study and take responsibility for the integrity of the data and the accuracy of the data analysis. Dr. Perlis is an associate editor for *JAMA Network Open* but was not involved in the editorial review or decision for this manuscript.

Dr. Perlis has received consulting fees from Burrage Capital, Genomind, Belle Artificial Intelligence, and Takeda. He holds equity in Psy Therapeutics, Belle Artificial Intelligence, and Circular Genomics. Dr. Edlow has received consulting fees from Mirvie, Inc. and research funding from Merck Pharmaceuticals, both outside of the work reported here. The other authors report no disclosures.

## Author Contributions

Andrea G. Edlow, MD MSc – planned analysis, drafted manuscript, revised manuscript

Victor M. Castro, MS – conducted data cleaning and analysis, revised manuscript

Lydia L. Shook, MD – revised manuscript

Sebastien Haneuse, Phd – revised manuscript, advised on data analysis

Anjali J. Kaimal, MD MAS – revised manuscript

Roy H. Perlis, MD MSc – planned analysis, drafted manuscript, revised manuscript, fed turtle

## Data Availability Statement

Data not available.

