## Supplemental Materials for "Sex-specific neurodevelopmental outcomes in offspring of mothers with SARS-CoV-2 in pregnancy: an electronic health records cohort"

**Table S1.** Frequency of individual developmental disorder ICD-10-CM codes occurring between delivery and 12 months in SARS-COV-2 exposed and unexposed mothers and within male and female offspring. (Cell counts below 5 are indicated with ‘<5’ per policy)

|  |  | Pregnancy SARS-COV-2 Negative | | | Pregnancy SARS-COV-2 Positive | | |
| --- | --- | --- | --- | --- | --- | --- | --- |
| ICD-10-CM code | ICD-10-CM description | All offspring | Male offspring | Female offspring | All offspring | Male offspring | Female offspring |
| F82 | Specific developmental disorder of motor function | 163 | 77 | 86 | 19 | 12 | 7 |
| F80.9 | Developmental disorder of speech and language, unspecified | 57 | 37 | 20 | <5 | <5 | <5 |
| F89 | Unspecified disorder of psychological development | 53 | 30 | 23 | <5 | <5 | <5 |
| F80.1 | Expressive language disorder | 34 | 26 | 8 | 0 | 0 | 0 |
| F88 | Other disorders of psychological development | 13 | 9 | <5 | <5 | 0 | <5 |
| F80.2 | Mixed receptive-expressive language disorder | <5 | <5 | <5 | <5 | <5 | 0 |
| F80.4 | Speech and language development delay due to hearing loss | <5 | <5 | 0 | 0 | 0 | 0 |

**Table S2.** Frequency of individual developmental disorder ICD-10-CM codes occurring between delivery and 18 months in SARS-COV-2 exposed and unexposed mothers and within male and female offspring. (Cell counts below 5 are indicated with ‘<5’ per policy)

|  |  | Pregnancy SARS-COV-2 Negative | | | Pregnancy SARS-COV-2 Positive | | |
| --- | --- | --- | --- | --- | --- | --- | --- |
| ICD-10-CM code | ICD-10-CM description | All offspring | Male offspring | Female offspring | All offspring | Male offspring | Female offspring |
| F80.1 | Expressive language disorder | 248 | 168 | 80 | 9 | 7 | <5 |
| F80.9 | Developmental disorder of speech and language, unspecified | 211 | 144 | 67 | 23 | 15 | 8 |
| F82 | Specific developmental disorder of motor function | 218 | 103 | 115 | 21 | 13 | 8 |
| F89 | Unspecified disorder of psychological development | 103 | 58 | 45 | <5 | <5 | <5 |
| F88 | Other disorders of psychological development | 17 | 11 | 6 | <5 | <5 | <5 |
| F80.2 | Mixed receptive-expressive language disorder | 13 | 9 | <5 | <5 | <5 | 0 |
| F80.89 | Other developmental disorders of speech and language | <5 | <5 | 0 | 0 | 0 | 0 |
| F80.4 | Speech and language development delay due to hearing loss | <5 | <5 | 0 | 0 | 0 | 0 |
| F80.0 | Phonological disorder | <5 | <5 | <5 | 0 | 0 | 0 |
| F81.9 | Developmental disorder of scholastic skills, unspecified | <5 | <5 | 0 | 0 | 0 | 0 |

**Figure S1.A**. Forest plot of developmental disorder outcome up to 12-months in SARS-COV-2 mothers with male offspring exact-matched to unexposed controls with male offspring. Cases were matched on maternal age, maternal race, maternal ethnicity, maternal insurance type, delivery hospital type and pre-term birth.

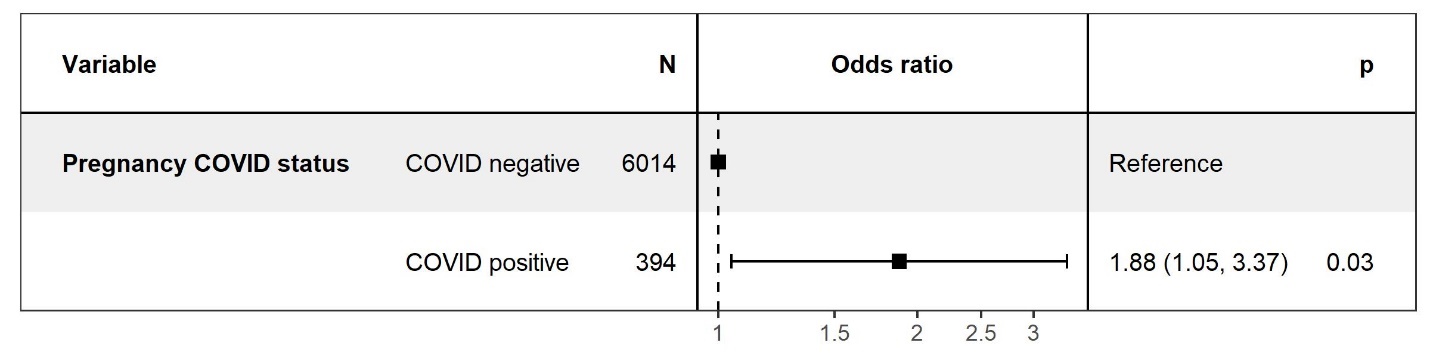

**Figure S1.B.** Forest plot of developmental disorder outcome up to 12-months in SARS-COV-2 mothers with female offspring exact-matched to unexposed controls with female offspring. Cases were matched on maternal age, maternal race, maternal ethnicity, maternal insurance type, delivery hospital type and pre-term birth.

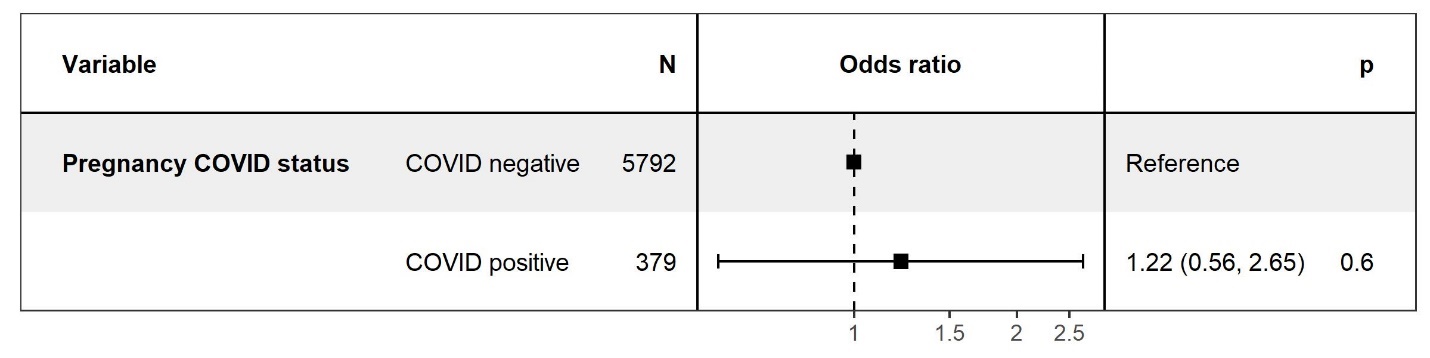

**Figure S1.C.** Forest plot of developmental disorder outcome up to 12-months in SARS-COV-2 mothers with all offspring exact-matched to unexposed controls. Cases were matched to controls on maternal age, maternal race, maternal ethnicity, maternal insurance type, delivery hospital type and pre-term birth.

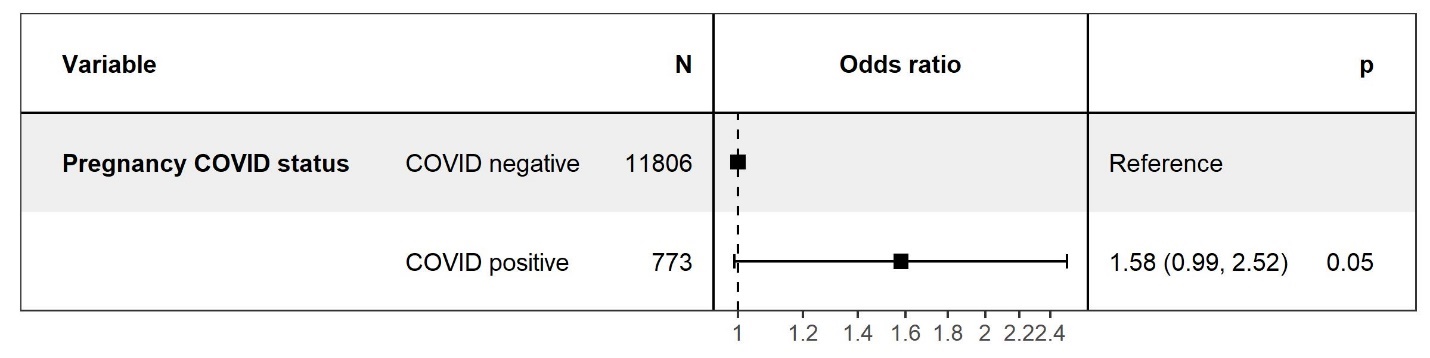

**Figure S2.A.** Forest plot of developmental disorder outcome up to 18-months in SARS-COV-2 mothers with male offspring exact-matched to unexposed controls. Cases were matched to controls on maternal age, maternal race, maternal ethnicity, maternal insurance type, delivery hospital type and pre-term birth.

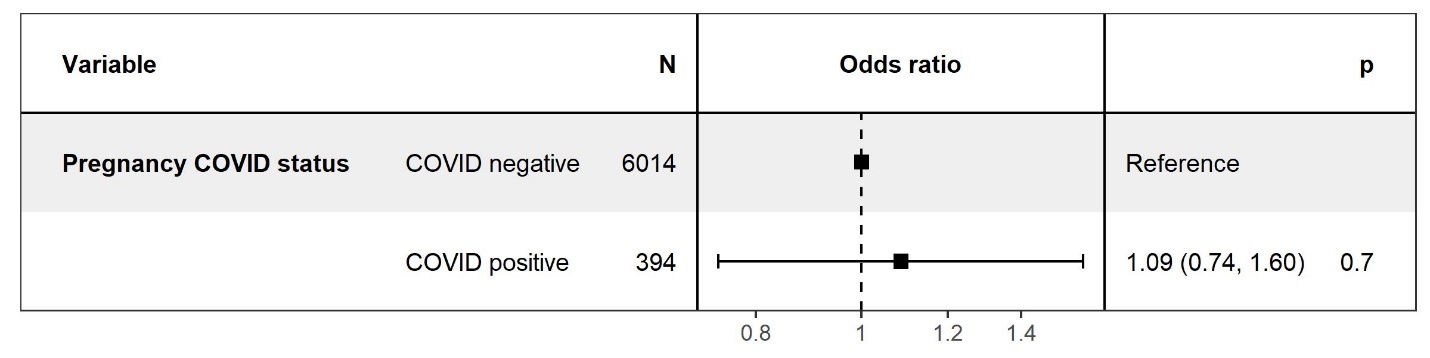

**Figure S2.B.** Forest plot of developmental disorder outcome up to 18-months in SARS-COV-2 mothers with female offspring exact-matched to unexposed controls. Cases were matched to controls on maternal age, maternal race, maternal ethnicity, maternal insurance type, delivery hospital type and pre-term birth.

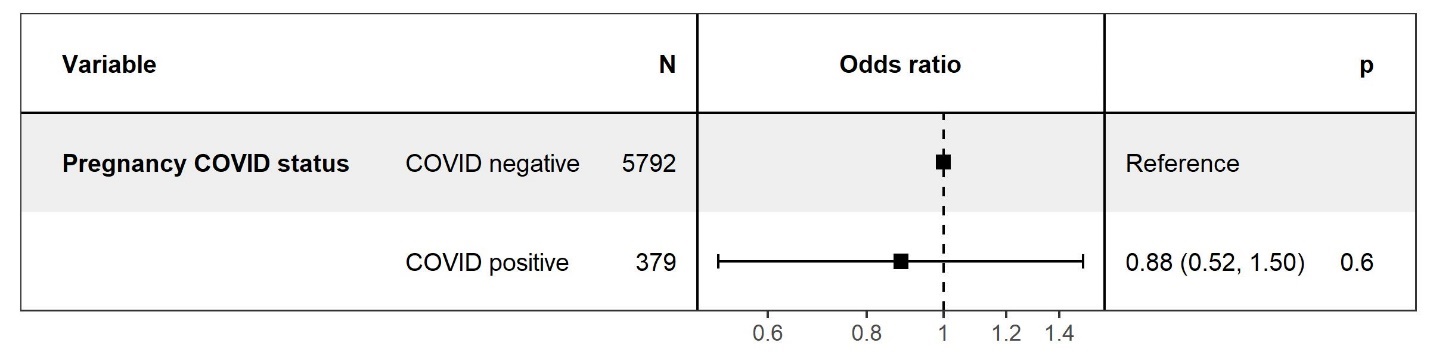

**Figure S2.C.** Forest plot of developmental disorder outcome up to 18-months in SARS-COV-2 mothers with all offspring exact-matched to unexposed controls. Cases were matched to controls on maternal age, maternal race, maternal ethnicity, maternal insurance type, delivery hospital type and pre-term birth.

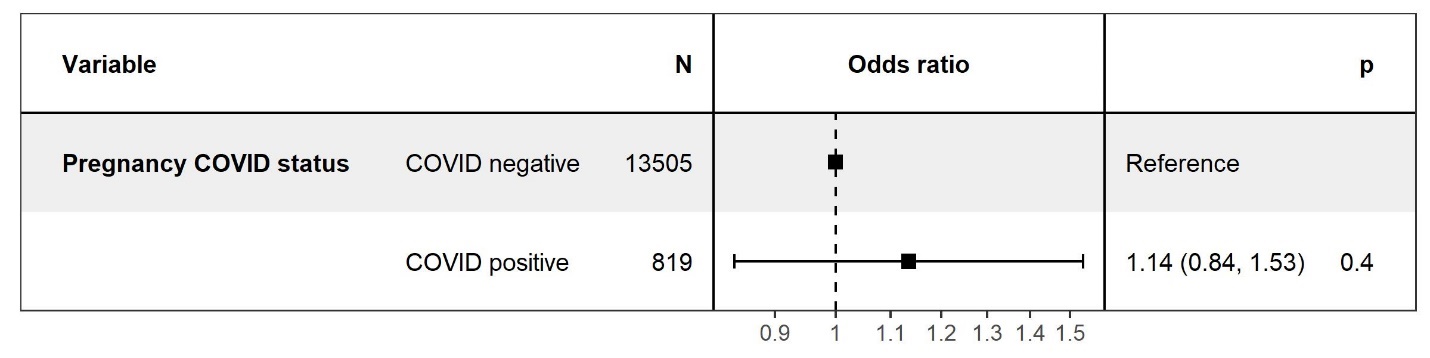

**Figure S3.A.** Logistic regression forest plot of developmental disorder outcome up to 12-months in male offspring stratified by maternal SARS-COV-2 exposure, maternal age, maternal race and ethnicity, maternal public insurance, and delivery hospital type (not stratified by pre-term birth)

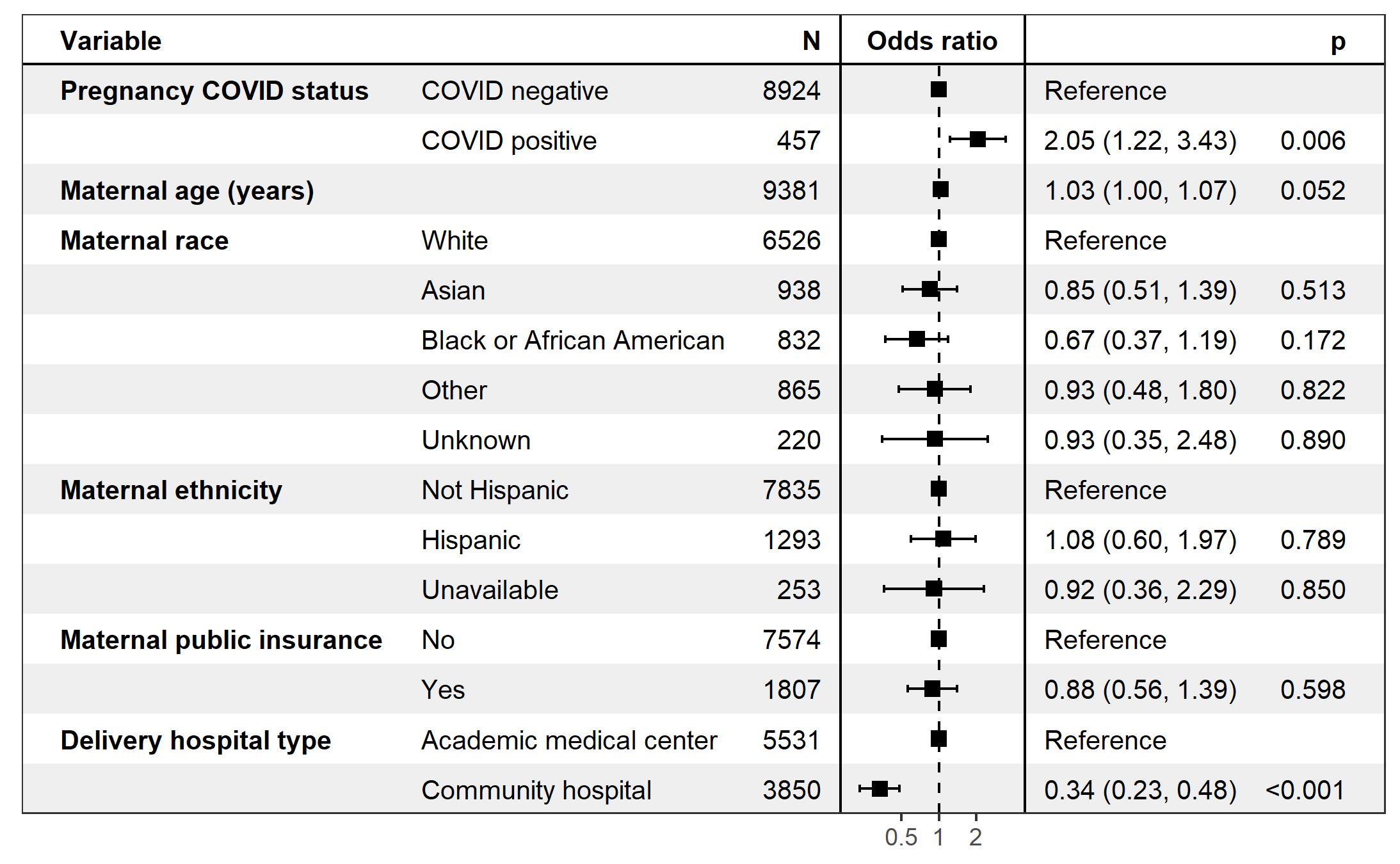

**Figure S3.B.** Logistic regression forest plot of developmental disorder outcome up to 12-months in female offspring stratified by maternal SARS-COV-2 exposure, maternal age, maternal race and ethnicity, maternal public insurance, and delivery hospital type (not stratified by pre-term birth)

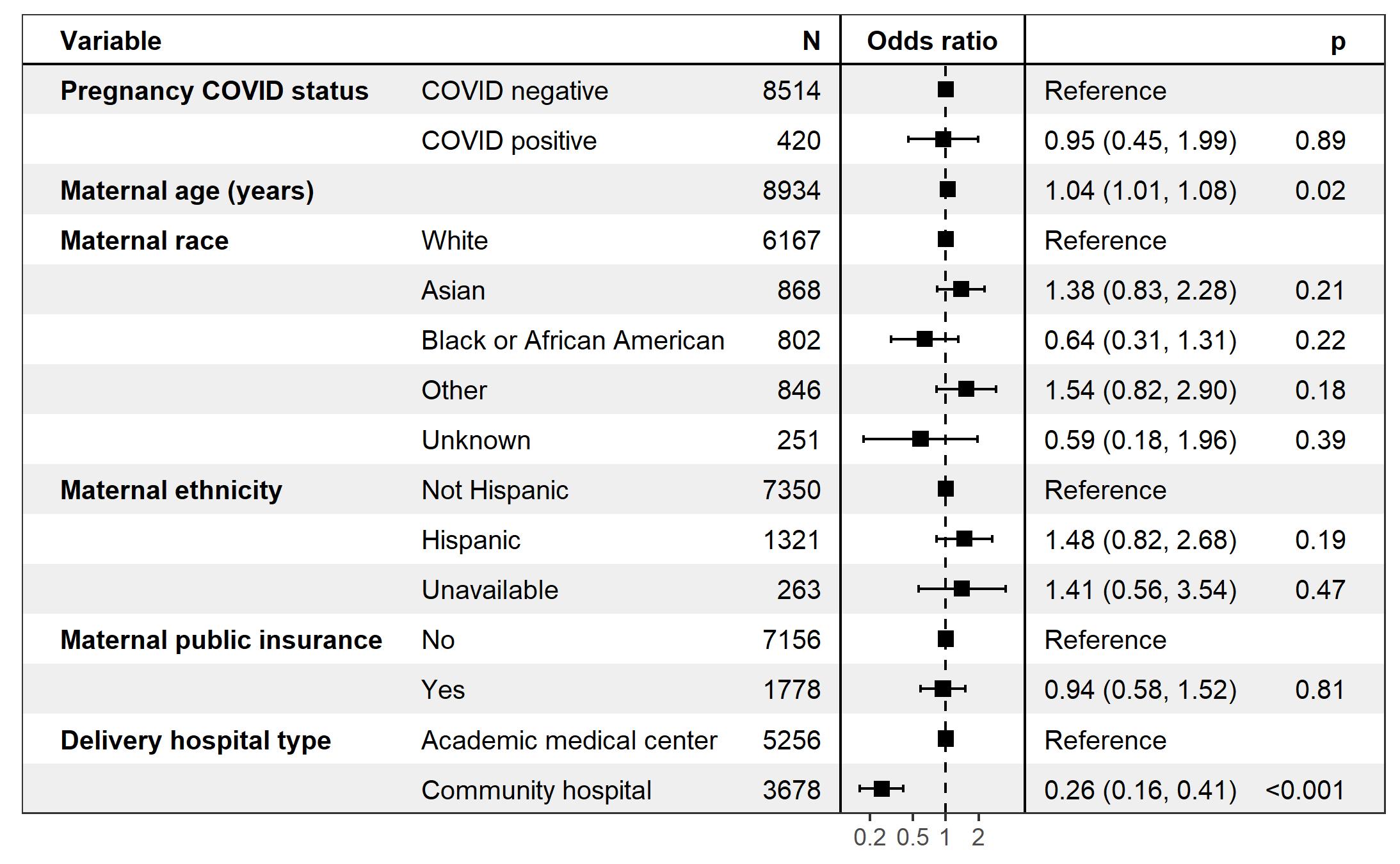

**Figure S3.C.** Logistic regression forest plot of developmental disorder outcome up to 12-months stratified by maternal SARS-COV-2 exposure, maternal age, maternal race and ethnicity, maternal public insurance, delivery hospital type and offspring sex (not stratified by pre-term birth)

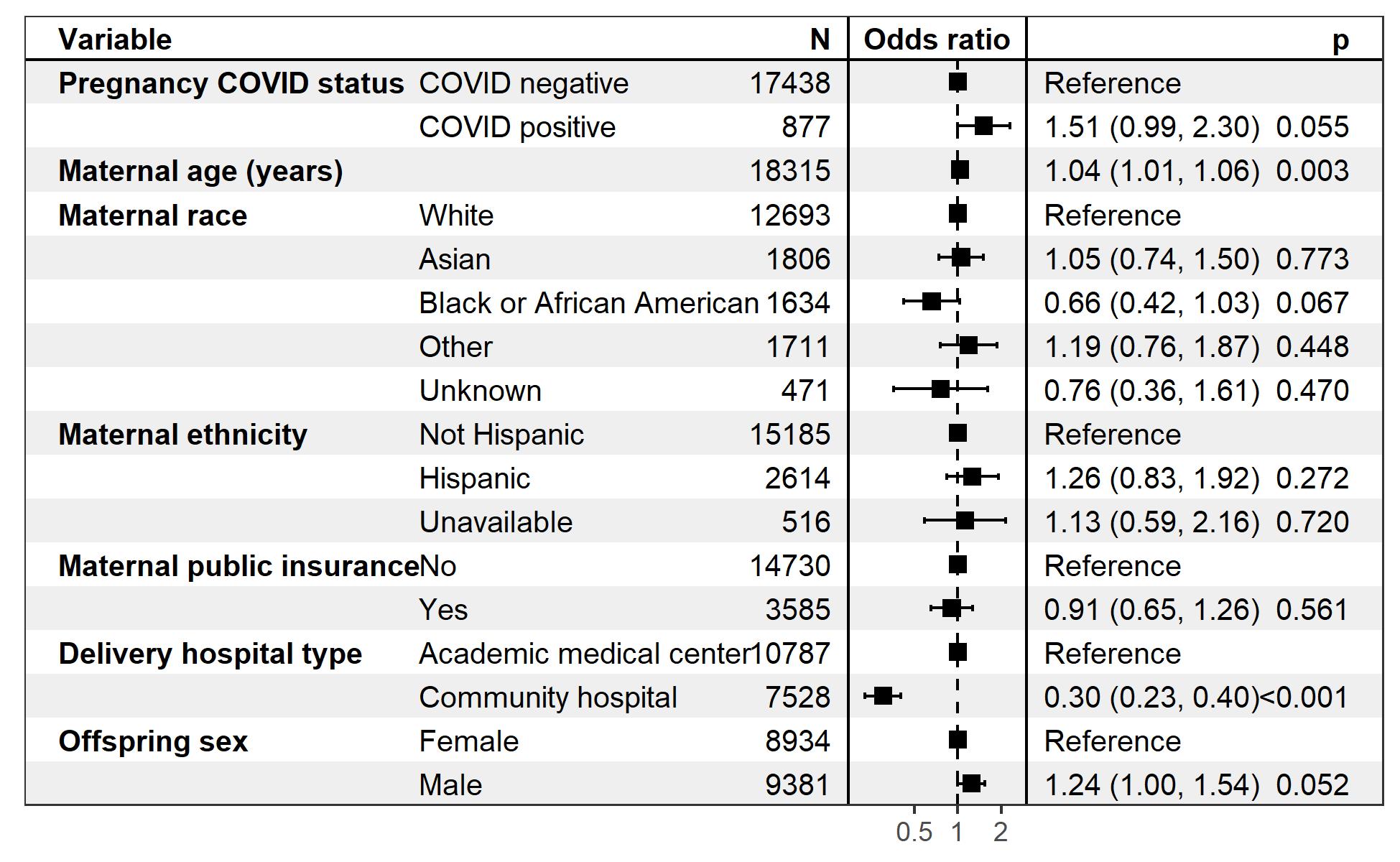

**Figure S4.A.** Logistic regression forest plot of developmental disorder outcome up to 18-months in male offspring stratified by maternal SARS-COV-2 exposure, maternal age, maternal race and ethnicity, maternal public insurance, and delivery hospital type (not stratified by pre-term birth)

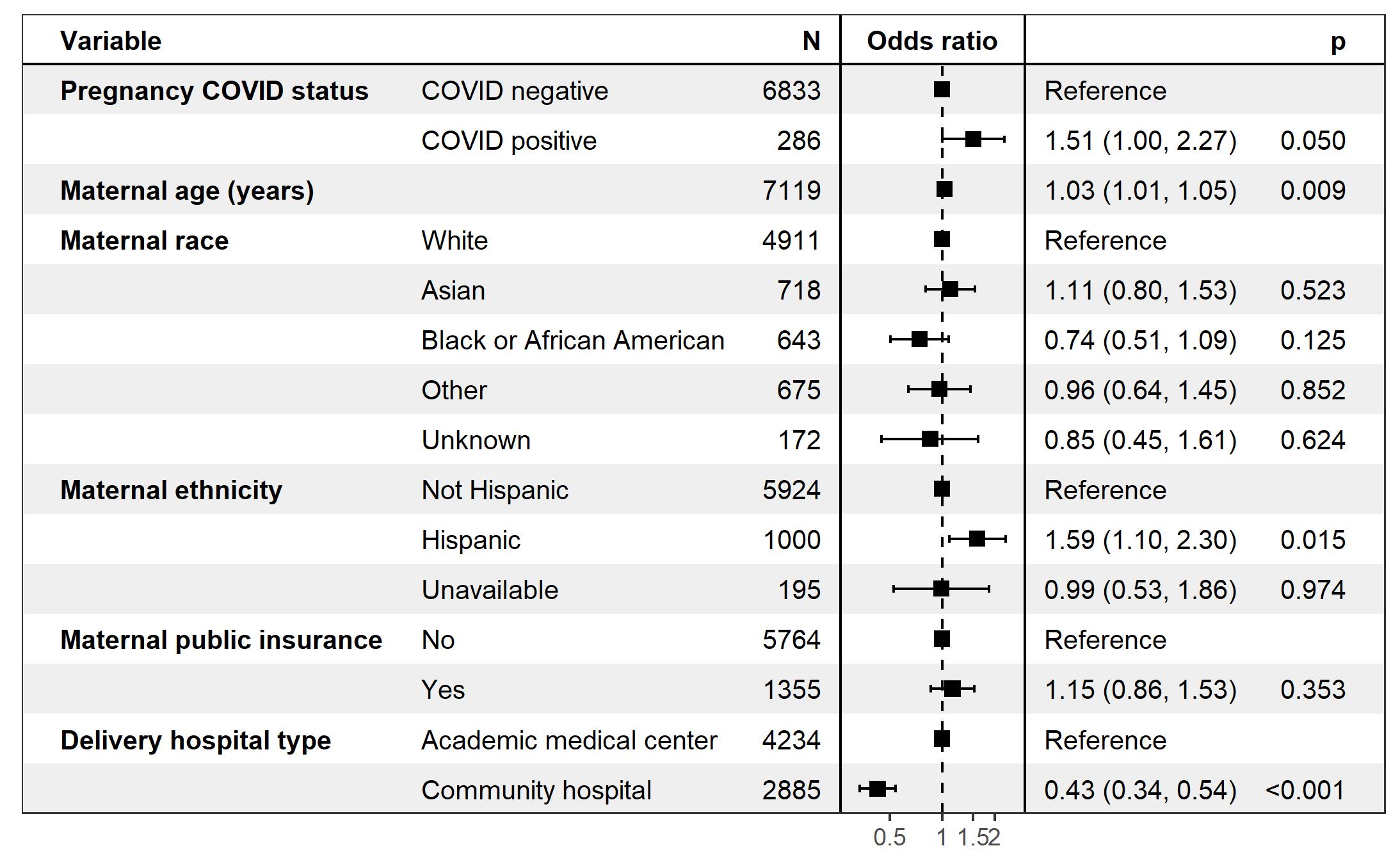

**Figure S4.B.** Logistic regression forest plot of developmental disorder outcome up to 18-months in female offspring stratified by maternal SARS-COV-2 exposure, maternal age, maternal race and ethnicity, maternal public insurance, and delivery hospital type (not stratified by pre-term birth)

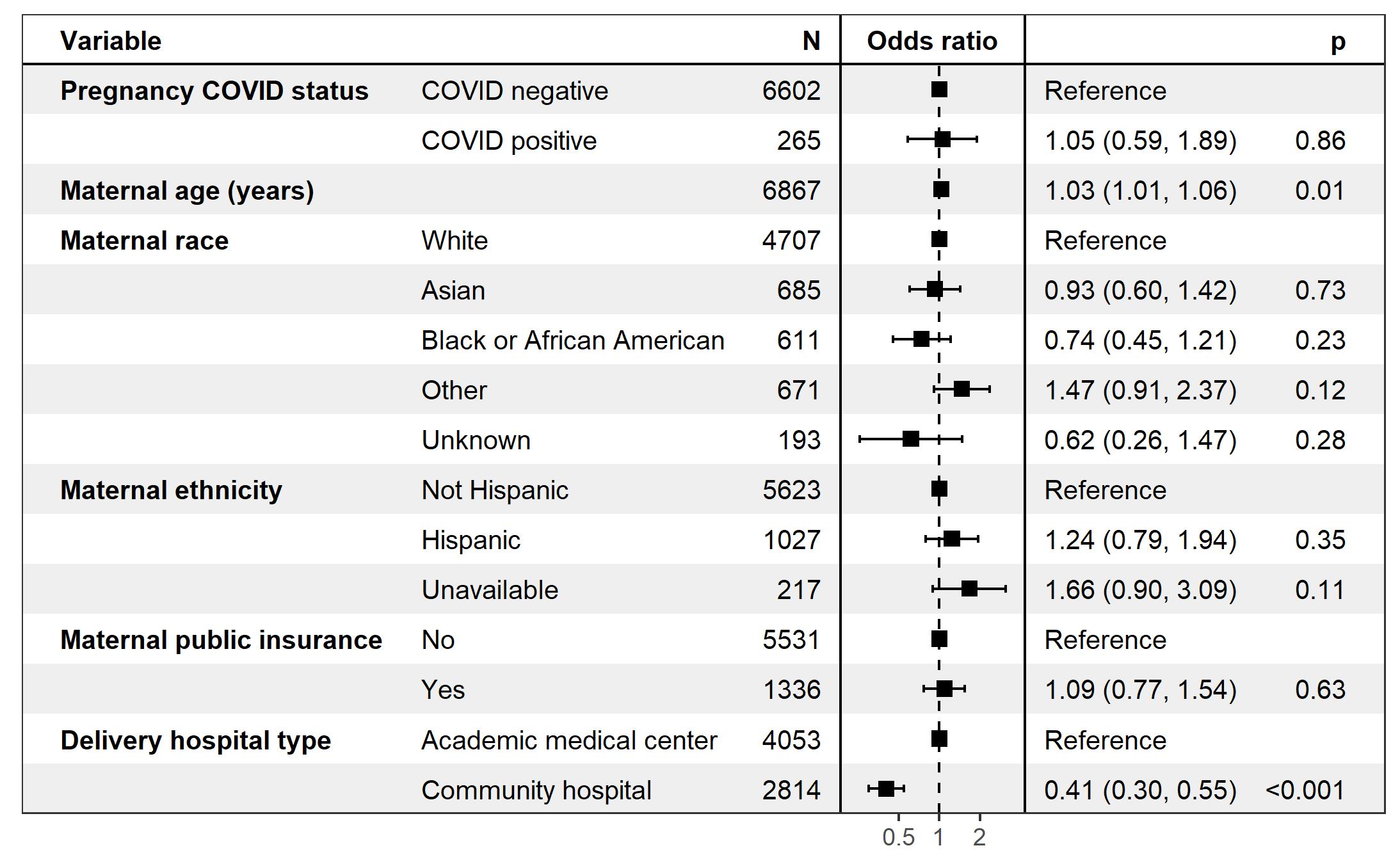

**Figure S4.C**. Logistic regression forest plot of developmental disorder outcome up to 18-months stratified by maternal SARS-COV-2 exposure, maternal age, maternal race and ethnicity, maternal public insurance, delivery hospital type and offspring sex (not stratified by pre-term birth)

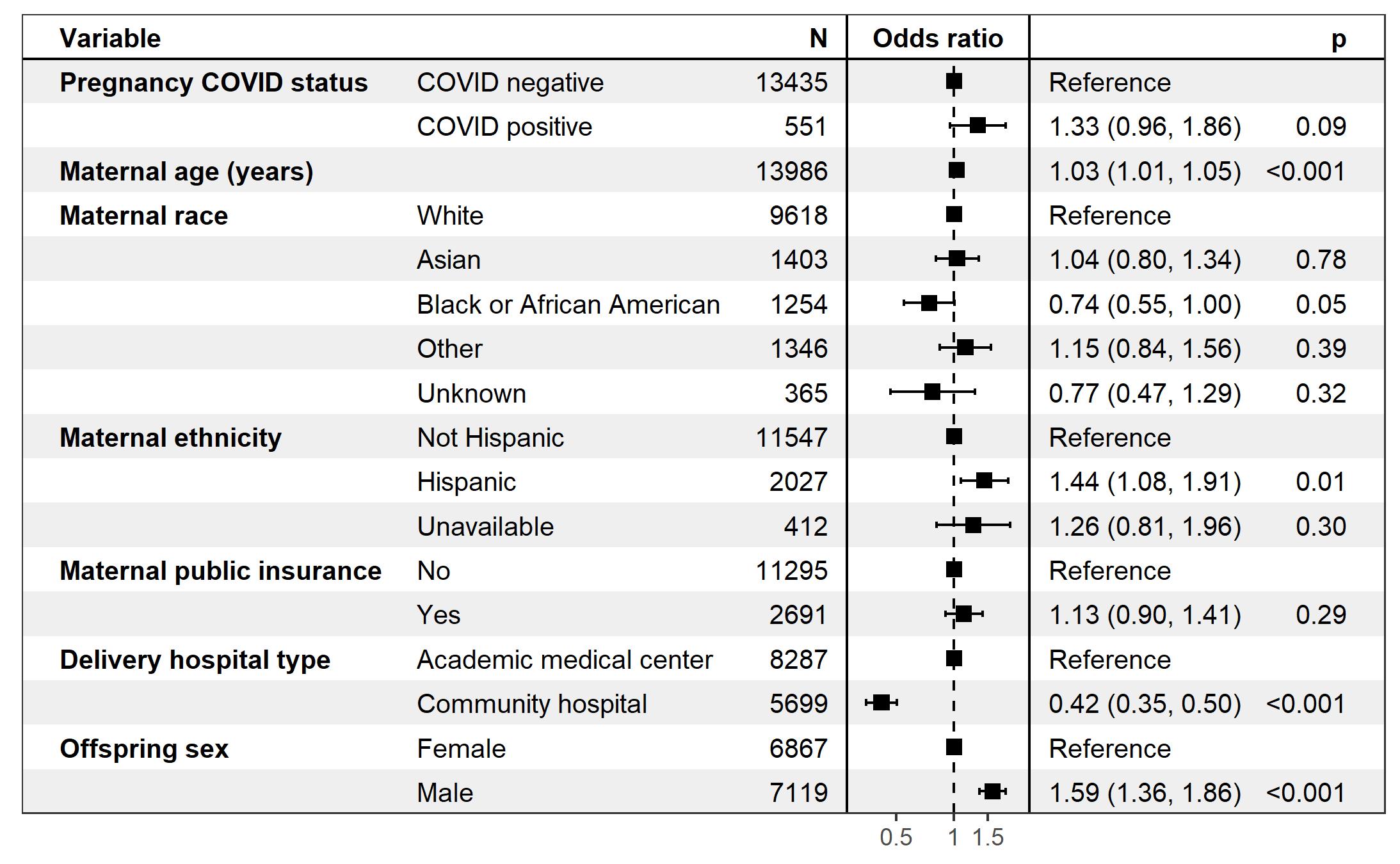

**Table S3.** Demographic and baseline clinical characteristic comparisons between births occurring in calendar year 2018 and births occurring during the COVID-19 pandemic study period (March 2020 - May 2021).

| Characteristic | Births in 2018 N=13,952 | Pandemic births N=18,323 | p-value^1^ |
| --- | --- | --- | --- |
| **Delivery hospital type, n (%)** |  |  | <0.001 |
| Academic medical center | 8,954 (64) | 10,789 (59) |  |
| Community hospital | 4,998 (36) | 7,534 (41) |  |
| **Maternal age, Mean (SD)** | 33.0 (4.9) | 33.0 (4.9) | 0.69 |
| **Maternal race, n (%)** |  |  | <0.001 |
| Asian | 1,544 (11) | 1,806 (9.9) |  |
| Black or African American | 1,320 (9.5) | 1,634 (8.9) |  |
| Other | 1,335 (9.6) | 1,711 (9.3) |  |
| Unknown | 430 (3.1) | 478 (2.6) |  |
| White | 9,323 (67) | 12,694 (69) |  |
| **Maternal ethnicity, n (%)** |  |  | <0.001 |
| Hispanic | 2,032 (15) | 2,614 (14) |  |
| Not Hispanic | 11,284 (81) | 15,185 (83) |  |
| Unavailable | 636 (4.6) | 516 (2.8) |  |
| Unknown | 0 | 8 |  |
| **Offspring gender, n (%)** |  |  | 0.21 |
| Female | 6,904 (49) | 8,938 (49) |  |
| Male | 7,048 (51) | 9,385 (51) |  |
| **Maternal public insurance, n (%)** | 2,714 (19) | 3,589 (20) | 0.76 |
| **Delivery method, n (%)** |  |  | 0.85 |
| C-Section | 4,596 (33) | 6,017 (33) |  |
| Vaginal | 9,356 (67) | 12,306 (67) |  |
| **Pre-term birth, n (%)** |  |  | 0.003 |
| Preterm | 1,555 (11) | 1,853 (10) |  |
| Term | 12,397 (89) | 16,470 (90) |  |
| **Delivery admission length of stay (days), Median (IQR)** | 3.00 (2.00 – 4.00) | 3.00 (2.00 – 4.00) | <0.001 |
| **Multiple births, n (%)** | 1,065 (7.6) | 1,165 (6.4) | <0.001 |
| ^1^Pearson's Chi-squared test; Wilcoxon rank sum test | | | |

**Figure S5.A**. Logistic regression forest plot of developmental disorder outcome up to 12-months in male offspring stratified by birth period (2018 vs pandemic study period), maternal age, maternal race and ethnicity, maternal public insurance, and delivery hospital type.

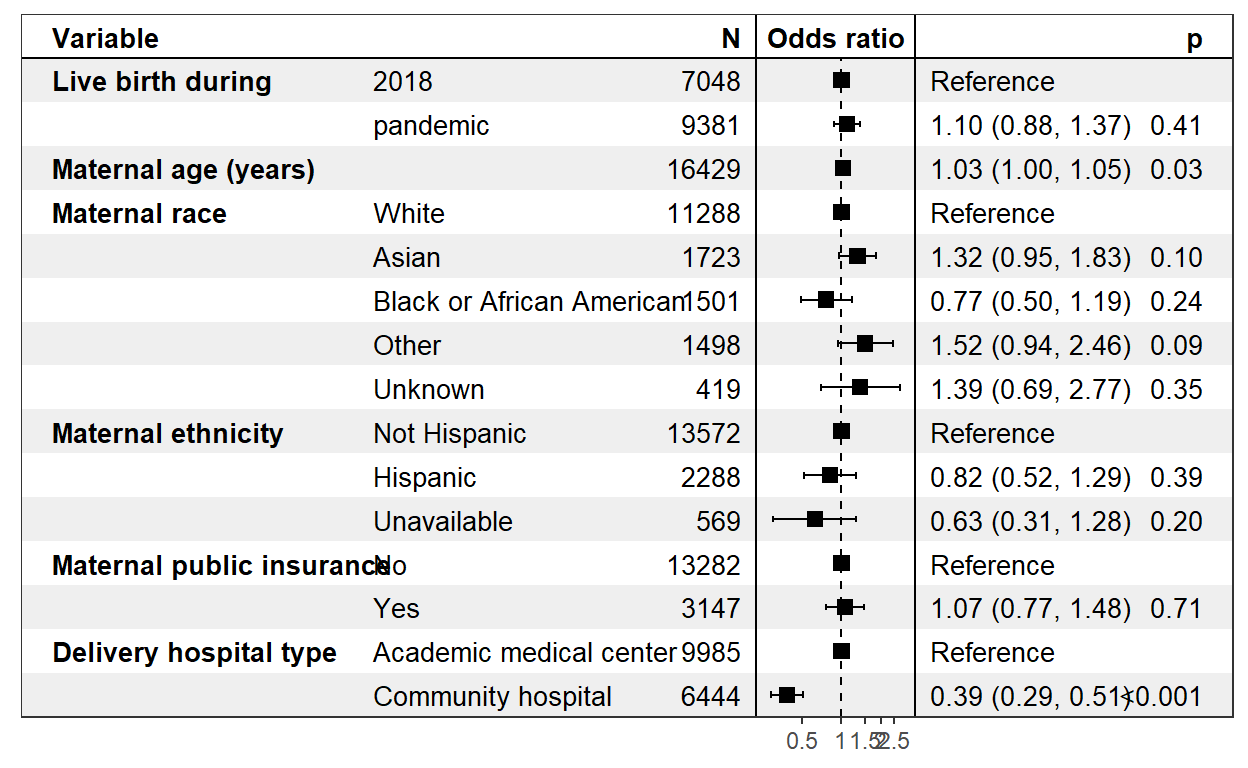

**Figure S5.B.** Logistic regression forest plot of developmental disorder outcome up to 12-months in female offspring stratified by birth period (2018 vs pandemic study period), maternal age, maternal race and ethnicity, maternal public insurance, and delivery hospital type.

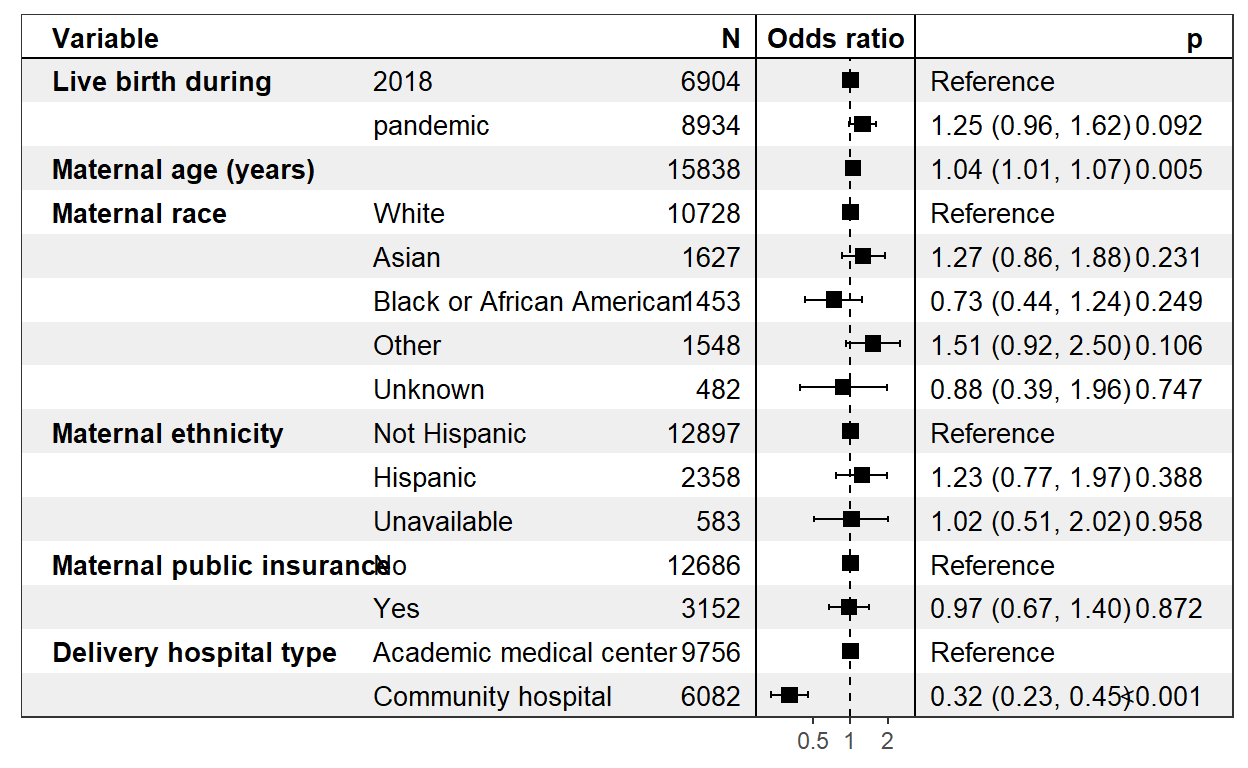

**Figure S5.C.** Logistic regression forest plot of developmental disorder outcome up to 12-months in all offspring stratified by birth period (2018 vs pandemic study period), maternal age, maternal race and ethnicity, maternal public insurance, and delivery hospital type.

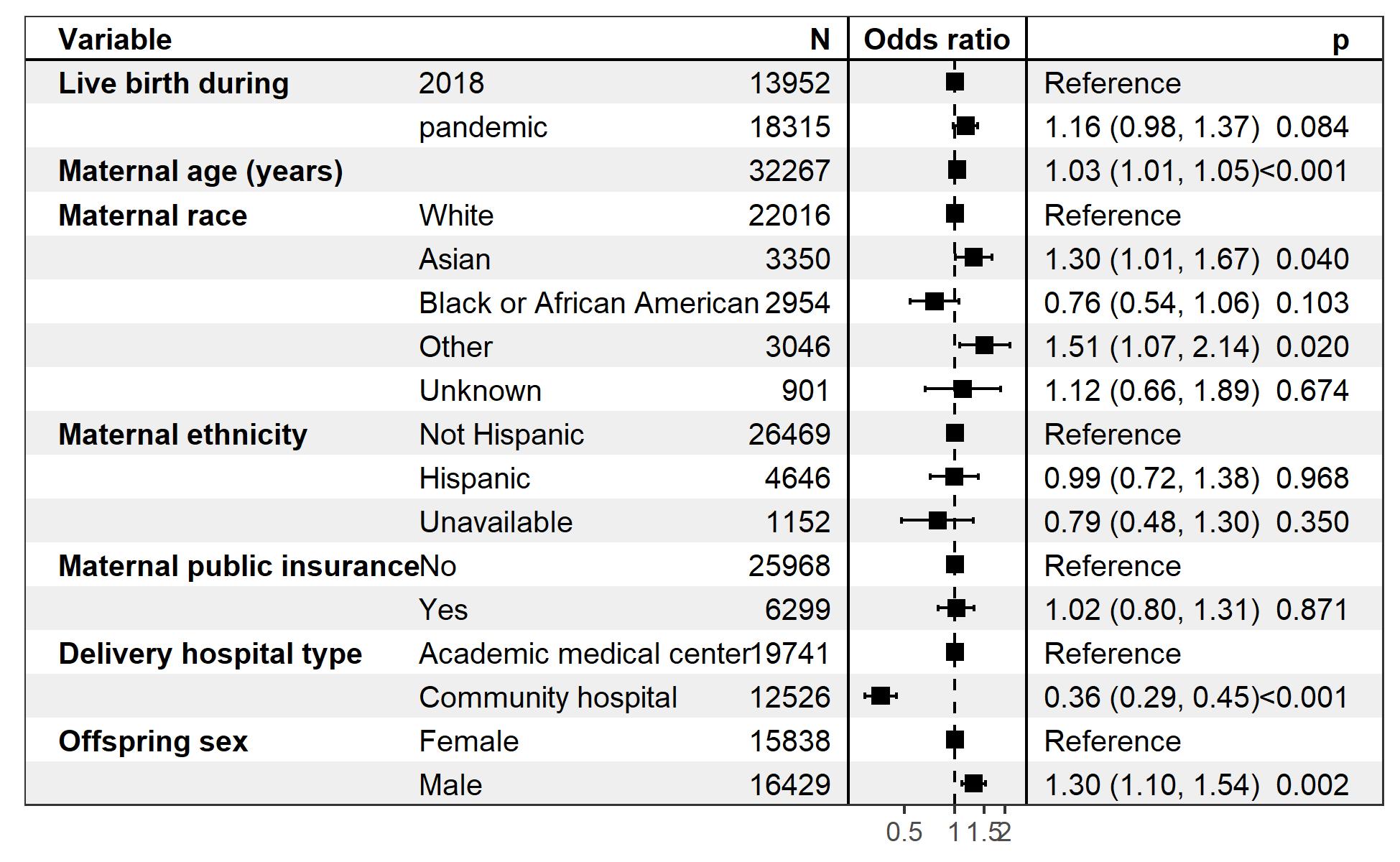

**Table S4.** Demographic and baseline clinical characteristic comparisons between births occurring in calendar year 2019 and births occurring during the COVID-19 pandemic study period (March 2020 - May 2021).

| Characteristic | **Births in 2019 N=15,068** | **Pandemic births N=18,323** | **p-value^1^** |
| --- | --- | --- | --- |
| **Delivery hospital type, n (%)** |  |  | <0.001 |
| Academic medical center | 9,564 (63) | 10,789 (59) |  |
| Community hospital | 5,504 (37) | 7,534 (41) |  |
| **Maternal age, Mean (SD)** | 33.1 (4.9) | 33.0 (4.9) | 0.60 |
| **Maternal race, n (%)** |  |  | <0.001 |
| Asian | 1,714 (11) | 1,806 (9.9) |  |
| Black or African American | 1,355 (9.0) | 1,634 (8.9) |  |
| Other | 1,499 (9.9) | 1,711 (9.3) |  |
| Unknown | 455 (3.0) | 478 (2.6) |  |
| White | 10,045 (67) | 12,694 (69) |  |
| **Maternal ethnicity, n (%)** |  |  | 0.14 |
| Hispanic | 2,260 (15) | 2,614 (14) |  |
| Not Hispanic | 12,365 (82) | 15,185 (83) |  |
| Unavailable | 438 (2.9) | 516 (2.8) |  |
| Unknown | 5 | 8 |  |
| **Offspring gender, n (%)** |  |  | 0.14 |
| Female | 7,472 (50) | 8,938 (49) |  |
| Male | 7,596 (50) | 9,385 (51) |  |
| **Maternal public insurance, n (%)** | 3,015 (20) | 3,589 (20) | 0.34 |
| **Delivery method, n (%)** |  |  | 0.94 |
| C-Section | 4,942 (33) | 6,017 (33) |  |
| Vaginal | 10,126 (67) | 12,306 (67) |  |
| **Pre-term birth, n (%)** |  |  | 0.007 |
| Preterm | 1,662 (11) | 1,853 (10) |  |
| Term | 13,406 (89) | 16,470 (90) |  |
| **Delivery admission length of stay (days), Median (IQR)** | 3.00 (2.00 – 4.00) | 3.00 (2.00 – 4.00) | <0.001 |
| **Multiple births, n (%)** | 1,076 (7.1) | 1,165 (6.4) | 0.004 |
| ^1^Pearson's Chi-squared test; Wilcoxon rank sum test | | | |

**Figure S6.A.** Logistic regression forest plot of developmental disorder outcome up to 12-months in male offspring stratified by birth period (2019 vs pandemic study period), maternal age, maternal race and ethnicity, maternal public insurance, and delivery hospital type.

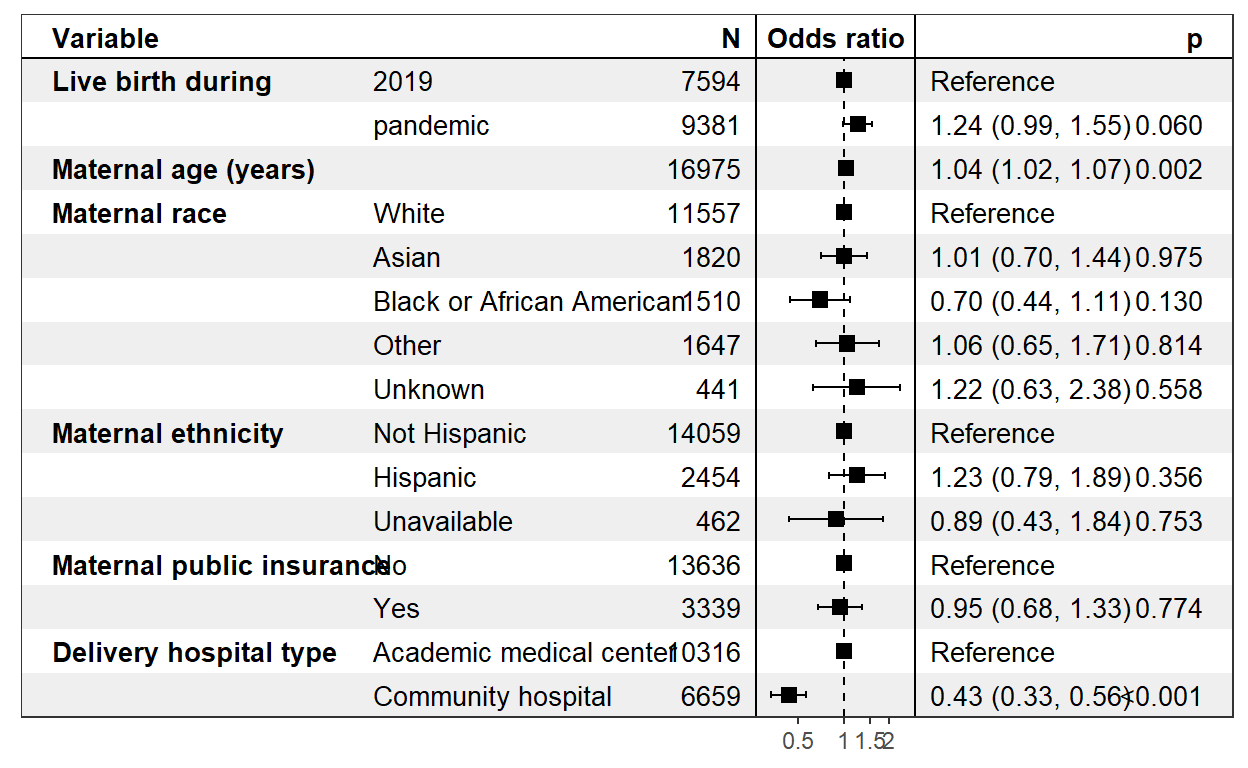

**Figure S6.B.** Logistic regression forest plot of developmental disorder outcome up to 12-months in female offspring stratified by birth period (2019 vs pandemic study period), maternal age, maternal race and ethnicity, maternal public insurance, and delivery hospital type.

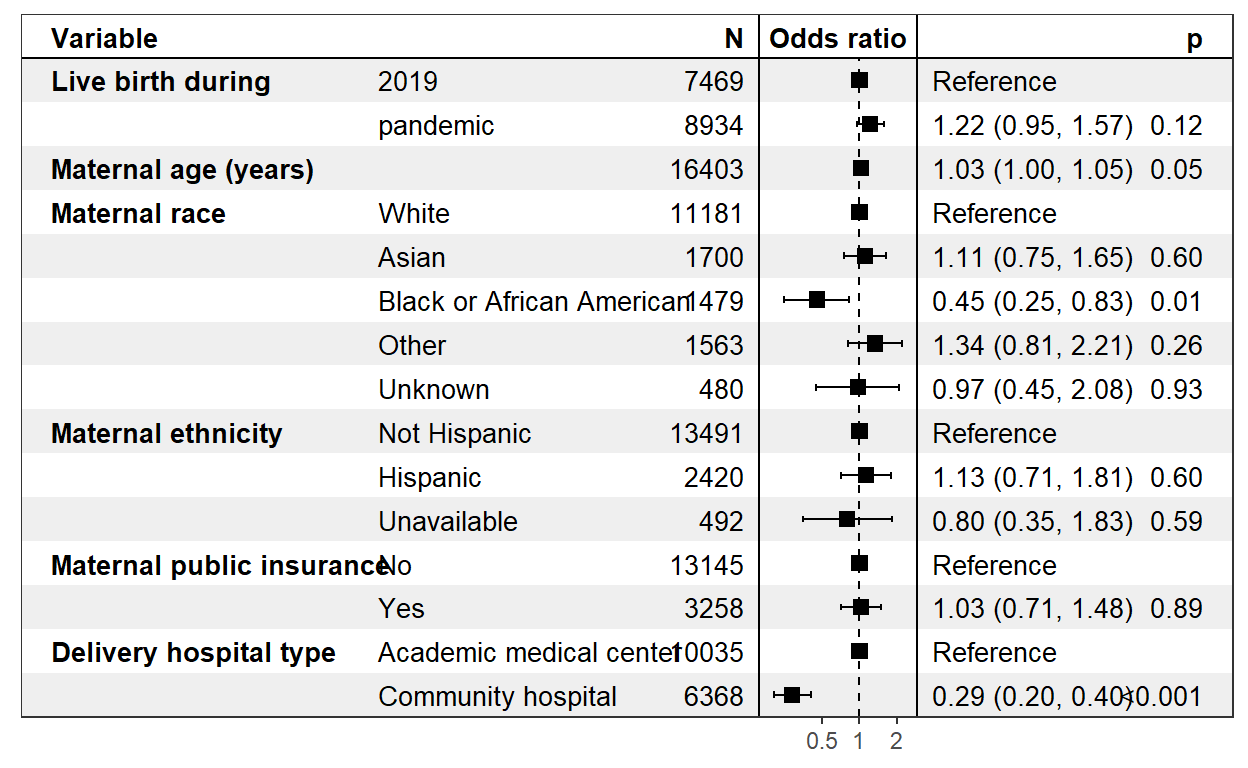

**Figure S6.C.** Logistic regression forest plot of developmental disorder outcome up to 12-months in all offspring stratified by birth period (2019 vs pandemic study period), maternal age, maternal race and ethnicity, maternal public insurance, offspring sex, and delivery hospital type.

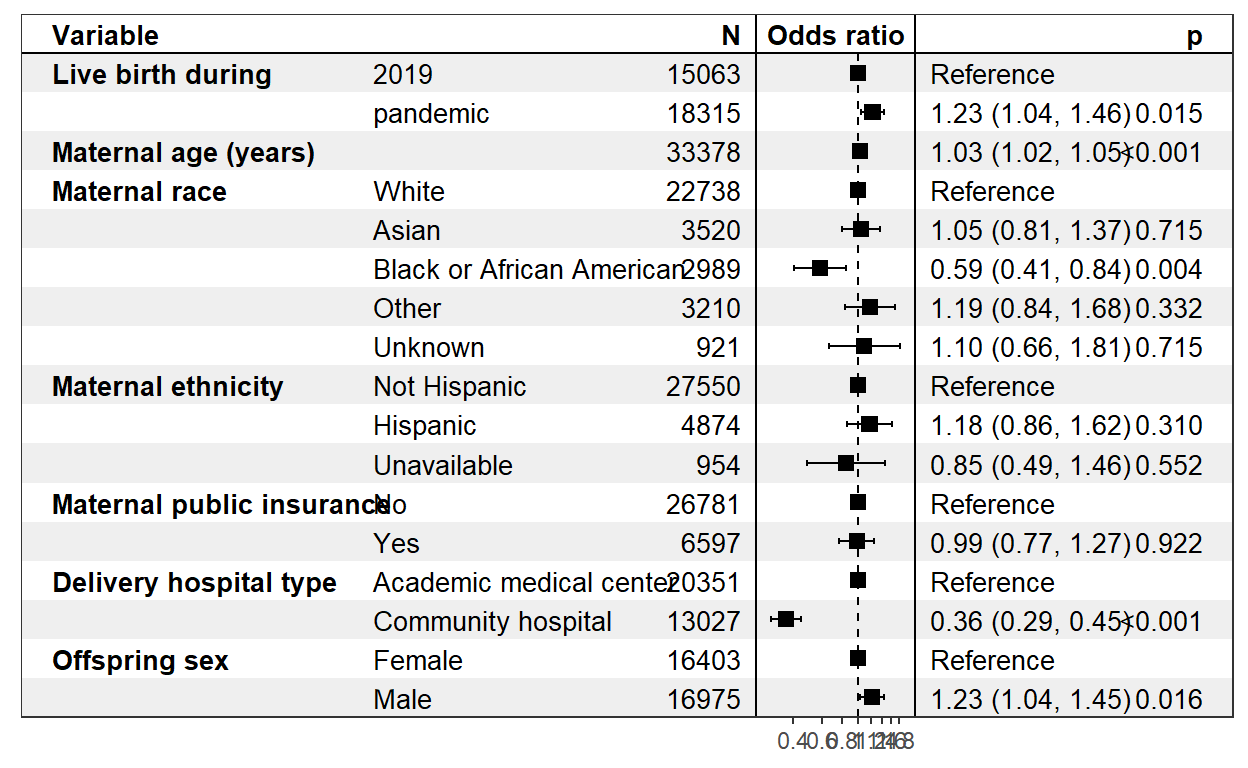

**Table S5.** Frequency of individual developmental disorder ICD-10-CM codes occurring between delivery and 12 months in births in 2018 and 2019 and within male and female offspring. (Cell counts below 5 are indicated with ‘<5’ per policy).

|  |  | Live births in 2018 | | | Live births in 2019 | | |
| --- | --- | --- | --- | --- | --- | --- | --- |
| ICD10CD | Description | All offspring | Male offspring | Female offspring | All offspring | Male offspring | Female offspring |
| F82 | Specific developmental disorder of motor function | 108 | 50 | 58 | 118 | 56 | 62 |
| F80.9 | Developmental disorder of speech and language, unspecified | 54 | 33 | 21 | 55 | 35 | 20 |
| F80.1 | Expressive language disorder | 35 | 25 | 10 | 39 | 23 | 16 |
| F89 | Unspecified disorder of psychological development | 23 | 16 | 7 | 23 | 15 | 8 |
| F88 | Other disorders of psychological development | 13 | 8 | 5 | 8 | 7 | <5 |
| F80.0 | Phonological disorder | <5 | 0 | <5 | <5 | <5 | 0 |
| F80.2 | Mixed receptive-expressive language disorder | 0 | 0 | 0 | <5 | 0 | <5 |
| F80.89 | Other developmental disorders of speech and language | <5 | <5 | 0 | <5 | <5 | 0 |
| F80.4 | Speech and language development delay due to hearing loss | <5 | <5 | <5 | <5 | <5 | 0 |
| F80.81 | Childhood onset fluency disorder | <5 | <5 | 0 | 0 | 0 | 0 |
